# Advancing Human Genetics Research and Drug Discovery through Exome Sequencing of the UK Biobank

**DOI:** 10.1101/2020.11.02.20222232

**Authors:** Joseph D. Szustakowski, Suganthi Balasubramanian, Ariella Sasson, Shareef Khalid, Paola G. Bronson, Erika Kvikstad, Emily Wong, Daren Liu, J. Wade Davis, Carolina Haefliger, A. Katrina Loomis, Rajesh Mikkilineni, Hyun Ji Noh, Samir Wadhawan, Xiaodong Bai, Alicia Hawes, Olga Krasheninina, Ricardo Ulloa, Alex Lopez, Erin N. Smith, Jeff Waring, Christopher D. Whelan, Ellen A. Tsai, John Overton, William Salerno, Howard Jacob, Sandor Szalma, Heiko Runz, Greg Hinkle, Paul Nioi, Slavé Petrovski, Melissa R. Miller, Aris Baras, Lyndon Mitnaul, Jeffrey G. Reid, on behalf of the UKB-ESC Research Team

## Abstract

The UK Biobank Exome Sequencing Consortium (UKB-ESC) is a unique private/public partnership between the UK Biobank and eight biopharma companies that will sequence the exomes of all ∼500,000 UK Biobank participants. Here we describe early results from the exome sequence data generated by this consortium for the first ∼200,000 UKB subjects and the key features of this project that enabled the UKB-ESC to come together and generate this data.

Exome sequencing data from the first 200,643 UKB enrollees are now accessible to the research community. Approximately 10M variants were observed within the targeted regions, including: 8,086,176 SNPs, 370,958 indels and 1,596,984 multi-allelic variants. Of the ∼8M variants observed, 84.5% are coding variants and include 2,139,318 (25.3%) synonymous, 4,549,694 (53.8%) missense, 453,733 (5.4%) predicted loss-of-function (LOF) variants (initiation codon loss, premature stop codons, stop codon loss, splicing and frameshift variants) affecting at least one coding transcript. This open access data provides a rich resource of coding variants for rare variant genetic studies, and is particularly valuable for drug discovery efforts that utilize rare, functionally consequential variants.

Over the past decade, the biopharma industry has increasingly leveraged human genetics as part of their drug discovery and development strategies. This shift was motivated by technical advances that enabled cost-effective human genetics research at scale, the emergence of electronic health records and biobanks, and a maturing understanding of how human genetics can increase the probability of successful drug development. Recognizing the need for large-scale human genetics data to drive drug discovery, and the unique value of the open data access policies and contribution terms of the UK Biobank, the UKB-ESC was formed. This precompetitive collaboration has further strengthened the ties between academia and industry and provided teams an unprecedented opportunity to interact with and learn from the wider research community.

## Why are drug developers interested in human genetics?

Developing novel therapies to address unmet medical needs is a resource-intensive challenge characterized by high attrition rates (Dimasi, Grabowski, & Hansen, 2016). While biopharmaceutical companies have recently improved overall R&D productivity and success rates for drug candidates in late stage clinical development, the probability of a drug candidate proceeding from Phase 1 clinical trials through approval and launch remains near 10%. Most clinical failures (approximately 75%) are attributable to safety concerns or lack of efficacy (Dowden & Munro, 2019).

The potential of human genetics to increase the likelihood of successful drug discovery has long been recognized, though largely in anecdotal form. Anti-*PCSK9* cholesterol-lowering drugs are a highly publicized example where human genetic evidence for a target contributed to technical and regulatory success, providing rationale for further investment (Furtado & Giugliano, 2020). Human genetics may also identify potential liability phenotypes associated with a target. For example, it is plausible that gastro-intestinal adverse events observed in clinical trials of DGAT1 inhibitors (Karlsson, Knutsson, & Eriksson, 2013; Meyers et al., 2015) could have been predicted based on the causal link between rare, highly penetrant *DGAT1* variants and congenital diarrheal disorder (Haas et al., 2012). Genetics has also proven its value in bringing more precision to drug development, particularly in the context of large trials. Stratification of patients to enrich for signal has shown success in *PCSK9* inhibitors (Sabatine et al., 2017; Schwartz et al., 2018), providing the ability to select genetically defined patient-populations. Table S1 enumerates a wide array of approved drugs with targets supported by human genetics evidence.

Recently, several studies systematically characterized the role of human genetics in drug discovery (Cook et al., 2014; King et al., 2019; Nelson et al., 2015). These retrospective analyses of drug development successes and failures suggest that drug-target pairs with human genetics evidence are at least twice as likely to reach approval as those without. Moreover, those success rates increase with the strength of evidence. Targets whose strongest evidence is a GWAS hit demonstrate approximately twice the probability of success as do targets lacking human genetics evidence, whereas those associated with rare Mendelian disorders are at least five times as likely to be successful. Conversely, drugs whose targets lack genetic association with a relevant indication are less likely to progress to market (King et al., 2019). Furthermore, genetic association with a non-relevant phenotype increases the likelihood of corresponding adverse events (Nguyen et al., 2019).

Building on these and similar experiences, several frameworks for the systematic evaluation of genetically motivated targets have emerged. For example, the allelic series model leverages the existence of multiple, independent genetic alterations in a gene of interest to assess its potential as a drug target. Genetic dose-response curves can be used to investigate the relationship between variants in the allelic series and phenotypes of interest to assess the potential for a tractable therapeutic window (Plenge, Scolnick, & Altshuler, 2013). In addition, several promising drug-development candidate targets with robust genetic evidence were identified based on associations between disease phenotypes, biomarker endo-phenotypes, and functionally consequential genetic variants. Recent examples include *HSD17B13* for chronic liver disease (Abul-Husn et al., 2018), *TYK2* for multiple autoimmune disorders (Dendrou et al., 2016; Diogo et al., 2015; Minegishi et al., 2006), *NRXN1* for neuropsychiatric disease (Noh et al. 2017), and *ASGR1* for cardiovascular disease (Nioi et al., 2016)

Large-scale biobanks have emerged to catalyze biomedical research, in part due to widespread adoption of electronic health records, and maturation of experimental and computational platforms capable of cost-effective population-scale sequencing and analysis. This creates a unique opportunity for the scientific community to build upon these foundations and accelerate the use of human genetics to inform drug discovery. Innovative public-private partnerships in the precompetitive space have demonstrated value for furthering this acceleration (Dolgin, 2019). Here, we describe one such effort – the UK Biobank Exome Sequencing Consortium (UKB-ESC) – focused on generating Whole Exome Sequencing (WES) data for all subjects (apprx. 500,000) in the UK Biobank (UKB).

## Why are drug developers interested in UK Biobank?

Nominating, evaluating, and prioritizing potential drug targets based on human genetics is data intensive. The most interesting novel insights are likely to come from rare or private functional genetic variants leading to highly penetrant gain or loss of protein function (M. I. McCarthy et al., 2008). These are most effectively discovered through direct sequencing of large populations, or those enriched for functionally important variation. Human genetic data alone, however, are insufficient. Comprehensive, consistent, longitudinal phenotypic data that can be linked to the genetic data are needed to explore the breadth of biological consequences of genetic variation. Essential phenotypes may include accurate disease diagnosis, molecular and cellular biomarkers, treatment and outcome information, imaging endpoints, self-reported conditions, and environmental and lifestyle data.

Beyond target identification, access to large-scale human genetic data linked to phenotypes enables a variety of other key opportunities for drug development efforts. In addition to direct human genetic discovery, these cohorts are incredibly valuable for “instant replication” of insights from other human genetics studies, prediction of potential side-effects, recall sub-studies, rapid validation of targets proposed by other means, and unbiased study of the natural history of rare monogenic diseases of interest to drug developers. Fifteen years ago, the Wellcome Trust Case Control Consortium shifted the field from candidate gene studies to genome-wide association studies (Burton et al., 2007). Now large biobanks are moving us even further in the unbiased spectrum by recruiting population-based cohorts without regard to disease status. The disease-agnostic approach and collection of a wide variety of data and specimens allows an immense number of hypotheses to be tested. The UKB is an exemplar of such data, a unique resource that provides a rich substrate for a broad spectrum of biomedical research (Bycroft et al., 2018).

The objective of the UKB-ESC is a comprehensive assessment of the protein-coding genetic variation in the half-million UKB participants. Enhancing this already phenotypically rich resource enables researchers to extend their genetic investigations to include rare and private variants not captured in existing UKB chip-based genotypes.

As the exome is enriched for variants that are most readily interpretable and actionable, this consortium has prioritized deep sequencing of the protein-coding portions of the genome. This is the highest value assay to add to the UKB genotype resource, pairing the rarest coding variation with the common and rare variation captured via chip genotyping and imputation. The speed of whole exome sequencing (WES) is enabling a rapid acceleration of actionable scientific discoveries. While WES is the assay of choice for Mendelian disease discovery (Y. Yang et al., 2013) and has provided important novel target discoveries (Abul-Husn et al., 2018), we are also enthusiastic for the arrival of the UKB whole-genome sequencing data (https://www.ukbiobank.ac.uk/2019/09/uk-biobank-leads-the-way-in-genetics-research-to-tackle-chronic-diseases/), and recognize the value of continued investment in growing the UKB resource.

## Building a successful large-scale drug-discovery collaboration in genomics: lessons learned

Large-scale collaborative projects have driven transformative scientific discoveries, particularly in genetics and genomics (Auton et al., 2015; Bycroft et al., 2018b; Dunham et al., 2012; Lander et al., 2001; S. McCarthy et al., 2016; Methé et al., 2012; “The Cancer Genome Atlas Program - National Cancer Institute,”). Such collaborations provide a unique opportunity to unify scientific communities by enabling a diversity of voices and perspectives to produce insights that would be inaccessible to smaller, more homogeneous groups. Historically, large-scale collaborative life science projects have been typically funded by governments and non-profits to engage academic groups. More recently, government agencies, non-profit organizations, and academic institutions have sought industry partners for big science projects (https://www.nih.gov/research-training/accelerating-medicines-partnership-ampwww.nih.gov/research-training/accelerating-medicines-partnership-amp, https://www.alzdiscovery.org/). The UKB-ESC seeks to further strengthen the ties between academia and industry through a precompetitive collaboration. This model is enabled by a project structure that engages all parties as scientific contributors and incentivizes industry investment in a sharable data resource. We expect the output of this partnership will catalyze novel scientific discoveries, accelerate the development of new therapies, and ultimately improve patient outcomes. As both the co-funders of and scientific teams analyzing the UKB WES data, we have found the following principles essential to the success of this effort.

### First and foremost, the UKB-ESC is uniquely enabled by the UKB open data access policy

The biomedical research community has already made great use of UKB data, as evidenced by the rapidly growing body of scientific literature (https://www.ukbiobank.ac.uk/published-papers/). The UKB data access model is valuable in how it enables researchers to openly publish and commercialize results from and derivatives of the UKB data and incentivizes contributions back to the resource. This virtuous cycle enables academia and industry to be engaged and benefit from a body of work larger than any one entity can achieve alone.

### Second, the UKB-ESC scale and scope will enable unique, valuable scientific discoveries

The hunt for actionable genetic variants (e.g. rare with profound functional consequences, common with informative phenotypic associations) depends on the scale of available data, in terms of both sample size and phenotypic characterization. Building large, diverse, and deeply characterized cohorts is an enormous multidisciplinary undertaking. This is particularly challenging in high-value phenotyping where different modalities (e.g. imaging, proteomics, EMR data extraction) require diverse expertise, and the data often require significant curation and processing prior to research use. The UKB excels at such tasks within a user-friendly framework attractive to both academic and industrial researchers, creating a roadmap for other projects with similar aspirations.

### Third, the UKB’s data access and contribution terms invites pre-competitive collaboration by industry partners

The generation of sequencing data at this scale requires a substantial investment of time, personnel, and financial resources (https://www.ukbiobank.ac.uk/2018/01/regeneron-announces-major-collaboration-to-exome-sequence-uk-biobank-genetic-data-more-quickly/). The UKB policy of providing a window of data exclusivity for data generators (also a common feature of large-scale academic collaborations) was essential to the business case for an investment of this scale. In addition to the scientific drivers described above, participation in a consortium and the exclusivity period both provide tangible benefits. While the competitive commercial interests of companies may seem to preclude cooperation, an appropriately managed collaboration mitigates individual risk to each partner and provides value larger than the sum of the individual investments. Moreover, the finite window of exclusivity helps focus efforts on scientific projects that are most likely to identify novel targets and accelerate therapeutic development.

### Finally, the value of engagement in a large pre-competitive industry collaborative project provides additional value to the participating institutions

Building expertise and functional excellence in important new areas is a key benefit to participating institutions—particularly in the rapidly transforming area of genetics research in support of drug discovery. In our experience, drug discovery teams typically have deep experience in areas such as disease biology, chemistry, and translational research. In contrast, the inclusion of human genetics expertise varies between companies and projects. This discrepancy is often the result of a lack of large-scale data sets that are relevant to specific therapeutic areas or drug development programs. We expect that the increasing availability of large, well-annotated genetics resources will lead to additional industry investment in genetics.

In summary, we have identified the following key features of an effective pre-competitive industry collaboration: Build a large dataset with rich phenotypic characterization, noting that participant recontact is highly valued for answering new questions that emerge from the data. Provide researchers the opportunity to derive both academic and/or commercial value from the data in an unencumbered way. Provide some exclusivity/first-mover advantage to build a compelling business justification for participation and financing of the project. Enable data access providing insight generation for key internal therapeutic-focused scientists and R&D stakeholders within the collaborating institutions. Provide opportunities for constructive engagement with academic partners, and low-friction data sharing terms and platforms to enable the broadest possible suite of research projects. We are hopeful others will follow the UKB model and build significant data collections designed to engage a wide variety of stakeholders.

## Characterization of the first 200,000 exome sequences from the UK Biobank

The release of the first ∼200,000 sequenced exomes represent a milestone in the availability of large-scale genomics data. We provide here an overview of the phenotypes and genotypes available as part of this resource. Table 1 includes a summary of the clinical characteristics of the first 50K sequenced individuals, the current 200K sequenced and overall 500K participants. Definitions of UKB phenotypes are provided in Van Hout et al, 2020. Table 2 summarizes the variants observed in the first 200,000 exomes sequenced from the UK Biobank. The targeted region covered 38,997,831 bases and coverage exceeds 20X on 95.6% of sites on average. Approximately 10M variants were observed within the targeted regions. These include: 8,086,176 SNPs, 370,958 indels and 1,596,984 multi-allelic variants. Of the ∼8M SNPs observed, 84.5% are coding variants and include 2,139,318 (25.3%) synonymous, 4,549,694 (53.8%) missense, 453,733 (5.4%) predicted loss-of-function (LOF) variants (initiation codon loss, premature stop codons, stop codon loss, splicing and frameshift variants) affecting at least one coding transcript. Analysis of allele frequencies revealed 98% of synonymous, 99% of missense and 99% of LOF (at least one transcript) variants with a minor allele frequency (MAF) less than 1%. On a per individual basis, we observed a median number of 8,586 synonymous, 7,724 missense and 202 LOF variants. Restricting our analysis to variants that affect canonical transcripts, we observe a median of 142 LOFs per individual. The numbers are largely similar to published exome studies (Karczewski et al., 2020; Van Hout et al., 2020). In addition, we also analyzed the increase in number of genes with heterozygous and homozygous LOFs with the increase in number of sequenced samples. Previously, 17,718 genes with heterozygous LOF variants were observed in the UKB 50,000 exomes data (Van Hout et al., 2020), therefore, is it not surprising that we only observe a modest increase in that number (18,045 genes) in 200K exomes. 789 genes with at least one homozygous LOF variant was reported in UKB 50,000 exomes data. The number of genes with homozygous LOFs still appears to be increasing with a projection of 1981 genes with at least one homozygous LOF variant in 500K (1492 genes with at least one homozygous LOF seen in UKB 200K, Table 3).

**Table 1:** Demographics and clinical characteristics comparing publicly released whole exome sequencing on 50K, and 200K subjects together with all UKB participants. Phenotypes were defined using a combination of IC10 codes together with self-reported verbal questionnaire. Values are expressed as median (1^st^ and 3^rd^ quartile). Data corresponds to UKB phenotypic release basket UKBB_41065 (March 2020). Subjects exclude participants withdrawn from UK Biobank as of September 2020.

|  | UKB 50K | UKB 200K | UKB 500k |
| --- | --- | --- | --- |
| Date data downloaded | March 2020 | March 2020 | March 2020 |
| Number of subjects | 49,898 | 200,633 | 502,504 |
| Female, n(%) | 27,207 (54.5) | 110,479 (55.1) | 273,378 (54.4) |
| Age at assessment, years | 58 (50-63) | 58 (50-63) | 58 (50-63) |
| Body mass index, kg/m2 | 26.6 (24-30) | 26.6 (24-30) | 26.7 (24-30) |
| Number of imaged participants (%) | 14,174 (28.4) | 23,818 (11.9) | 44,703 (8.9) |
| Number of current/past smokers, n(%) | 22,327 (44.7) | 89,391 (44.6) | 226,901 (45.2) |
| Townsend Deprivation Index | -2.0 (-3.6,0.6) | -2.2 ( -3.7, 0.5) | -2.1 (-3.6, 0.6) |
| Inpatient ICD10 codes per patient | 8 | 8 | 8 |
| Patients with >=1 ICD10 diagnoses, n(%) | 43,345 (86.9) | 164,452 (82.0) | 410,310 (81.7) |
| **Genetic Ancestry Assignment** |  |  |  |
| African (%) | 2.0 | 1.6 | 1.6 |
| East Asian (%) | 0.35 | 0.32 | 0.31 |
| European (%) | 93.3 | 93.7 | 94.0 |
| **Enhanced measures** |  |  |  |
| Hearing test, n(%) | 40,761 (81.7) | 84,484 (42.1) | 171,826 (34.2) |
| Pulse Rate, n(%) | 40,495 (81.2) | 83,974 (41.9) | 170,743 (34.0) |
| Visual Acuity Measured, n(%) eids | 39,551 (79.3) | 65,248 (32.5) | 117,672 (23.4) |
| IOP measured (left),n(%) | 37,889 (75.9) | 62,213 (31.0) | 111,926 (22.3) |
| Autorefracton, n(%) eids | 39,520 (79.2) | 64,807 (32.3) | 116,365 (23.2) |
| Retinal OCT, n(%) eids | 38,371 (76.9) | 51,286 (25.6) | 78,888 (15.7) |
| ECG at rest, n(%) | 12,331 (24.7) | 15,953 (8.0) | 24,923 (5.0) |
| Cognitive Function, n(%) | 17,340 (34.8) | 53,039 (26.4) | 123,614 (24.6) |
| Digestive Health, n(%) | 23,922 (47.9) | 74,417 (37.1) | 174,760 (34.8) |
| Physical Activity Measurement, n(%) | 15,002 (30.1) | 44,885 (22.4) | 103,684 (20.6) |
| **Cardiometabolic phenotypes** |  |  |  |
| Coronary Disease, n(%) | 4,356 (8.7) | 16,224 (8.1) | 45,450 (9.0) |
| Heart Failure, n(%) | 707 (1.4) | 3,009 (1.5) | 8,939 (1.8) |
| Type 2 Diabetes, n(%) | 2,817 (5.6) | 11,150 (5.6) | 29,266 (5.8) |
| **Respiratory and immunological phenotypes** |  |  |  |
| Asthma, n(%) | 8,471 (17.0) | 28,491(14.2) | 70,969 (14.1) |
| COPD, n(%) | 1,481 (3.0) | 5,135 (2.6) | 14,688 (2.9) |
| Rheumatoid Arthritis, n(%) | 951 (1.9) | 3,802 (1.9) | 9,685 (1.9) |
| Inflammatory Bowel Disease n(%) | 650 (1.3) | 2,616 (1.3) | 6,851 (1.4) |
| **Neurodegenerative phenotypes** |  |  |  |
| Alzheimer's Disease, n(%) | 47 (0.09) | 352 (0.18) | 933 (0.19) |
| Parkinson's Disease, n(%) | 131 (0.26) | 721 (0.36) | 1,938 (0.39) |
| Multiple Sclerosis, n(%) | 163 (0.33) | 684 (0.34) | 1,766 (0.35) |
| Myasthenia Gravis, n(%) | 23 (0.05) | 129 (0.06) | 300 (0.06) |

**Table 2:** Summary statistics for variants in publicly released 200K exomes. Counts of autosomal variants observed across all individuals by type/functional class for all and for MAF<1%. The number of bases targeted for capture by the exome capture reagent was n=38,997,831. Median count and interquartile range (IQR) per individual for all variants, and for MAF<1%. Counts restricted to WES targeted regions. Variant annotation details are included in the Methods section.

| Variants in WES |  |  | Median Per Participant (IQR) |  |
| --- | --- | --- | --- | --- |
| n=200,643 Participants | # Variants | # Variants<br>MAF < 1% | # Variants | # Variants<br>MAF < 1% |
| Total | 16,846,188 | 16,709,690 | 44,719 (567) | 3,050 (107) |
| Targeted Regions | 8,457,134 | 8,395,913 | 21,380 (240) | 1,466 (50) |
| **Variant Type** |  |  |  |  |
| SNVs | 8,086,176 | 8,026,702 | 20,817 (234) | 1,403 (49) |
| Indels | 370,958 | 369,211 | 563 (23) | 64 (8) |
| Multi Allelic Sites | 1,596,984 | 1,585,412 | 3768 (68) | 223 (19) |
| **Functional Prediction** |  |  |  |  |
| Synonymous | 2,139,318 | 2,115,702 | 8,586 (114) | 457 (24) |
| Missense | 4,549,694 | 4,526,159 | 7,724 (115) | 677 (32) |
| LOF (any transcript) | 453,733 | 453,007 | 202 (15) | 31 (7) |
| LOF (strict) | 381,717 | 381,232 | 142 (12) | 22 (6) |

**Table 3:** Observed number of heterozygous and homozygous carriers of LOF variants in ∼200K exomes and projections for the number expected in 500K. Predicted number of genes in larger WES sample sizes from existing 200K WES data estimated based on the methodology described in Van Hout et al, 2020. The number of autosomal genes with at least 1, 5, 10, etc. heterozygous and homozygous LOF carriers passing QC filters described in Table 2 for 189,698 UKB participants of European ancestry with WES and predicted number of genes with N heterozygous and homozygous LOF carriers in 500k.

| LOF (AAF<1%) | 189,698 individuals | 500,000 individuals |
| --- | --- | --- |
| Minimum number of carriers (N) | Observed number of genes with at least N heterozygous carriers | Predicted number of genes with at least N heterozygous carriers |
| 1 | 18045 | 18414 |
| 5 | 17411 | 17968 |
| 10 | 16556 | 17514 |
| 25 | 14204 | 16384 |
| 50 | 11380 | 14853 |
| 100 | 7830 | 12475 |
| 250 | 3824 | 8065 |
| 500 | 1905 | 4711 |
| 1000 | 914 | 2449 |
| 2500 | 238 | 924 |
| 5000 | 18 | 394 |
| 10000 | 1 | 57 |
| Minimum number of Carriers | Observed number of genes with at least N homozygotes | Predicted number of genes with at least N homozygotes |
| 1 | 1492 | 1981 |
| 5 | 528 | 954 |
| 10 | 193 | 627 |
| 25 | 20 | 199 |
| 50 | 2 | 42 |
| 100 | 0 | 5 |
| 250 | 0 | 0 |
| 500 | 0 | 0 |
| 1000 | 0 | 0 |
| 2500 | 0 | 0 |
| 5000 | 0 | 0 |
| 10000 | 0 | 0 |

## Summary and closing thoughts

Transforming healthcare with data is at the heart of the UKB-ESC. We are proud of our contribution to the scientific community, enthusiastic about making these data broadly available, and excited to see what the future holds in terms of discoveries from and contributions to the UKB and UKB-ESC resources, particularly as we gain additional insight into functionally consequential variants that can meaningfully inform drug development. Data and methods developed by the UKB-ESC have already contributed to several publications (Liu et al., 2020).

With the UKB-ESC exome sequencing nearly complete, we hope the key features of this collaboration will be adopted as the preferred model for similar projects in the future. As an example, this pre-competitive industry collaboration model could be leveraged to generate complementary deep phenotyping data for UKB subjects and open entirely new avenues of scientific research. We expect the value of the WES data will be enhanced by layering deeper and richer phenotypes, which can provide important insights into disease biology, characterize response to marketed therapies, and identify novel target identification. Thoughtfully designed projects that maximize the engagement of academia, non-profits, and industry will yield valuable data resources and scientific insights that accelerate drug discovery and personalized healthcare.

There is no more important scientific challenge than developing new therapies to treat unmet medical needs. Large-scale collaborations such as the UKB-ESC play an essential role by generating unique, accessible resources that can be used by a diverse community of researchers to address questions critical to advancing human health.

## Data Availability

All data is avaiable through the UK Biobank (https://www.ukbiobank.ac.uk/) to bona fide researchers who have submitted compliant research applications.

## Data accessibility

UK Biobank aims to encourage and provide as wide access as possible to its data and samples for health-related research in the public interest by all bona fide researchers from the academic, charity, public, and commercial sectors, both in the UK and internationally, without preferential access for any user. UK Biobank’s publicly available Data Showcase (http://biobank.ndph.ox.ac.uk/showcase/) presents the univariate distributions and methods used for collection of all the variables available for health-related research, enabling potential research users to explore what data are available and plan research applications.

All researchers who wish to access the resource must register with UK Biobank via its online Access Management System (AMS) (https://bbams.ndph.ox.ac.uk/ams/). Once approved, researchers may apply (via the AMS) to access the resource for specific, well-defined research projects. At the time of publication, over 16,500 researchers were registered with UK Biobank and over 2,000 research applications were approved (see www.ukbiobank.ac.uk/approved-research/ for a summary of research that is currently underway).

## Acknowledgements

The authors would like to thank everyone who made this work possible, particularly the UK Biobank team, their funders, the dedicated professionals from the member institutions who contributed to and supported this work, and most especially the UK Biobank participants, without whom this research would not be possible. This research has been conducted using the UK Biobank Resource under application number 26041.

## Methods

### OQFE Protocol

The OQFE protocol maps raw reads (FASTQ) with BWA-MEM to the GRCh38 reference in a deterministic manner, retaining all supplementary alignments. Mate tags are added with samblaster as specified in the FE protocol (Regier et al., 2018). OQFE CRAMs contain all reads from the input FASTQs and meet all FE tag specifications. Duplicate reads are then marked with Picard 2.21.2, which resolves a known issue with the FE version of Picard (2.4.1), in which the representative read in a set of duplicate reads can depend on the sequence input order, potentially resulting in an order-dependent set of supplementary alignment duplicates. The final OQFE CRAM is compressed with samtools, without any quality score recalibration or binning.

### OQFE mapping and variant calling with UK Biobank exome sequencing

The OQFE protocol was applied to more than 200,000 UK Biobank (UKB 200K) exome samples with the OQFE pipeline (https://hub.docker.com/r/dnanexus/oqfe). Variants were called on each CRAM with DeepVariant 0.10.0 (Poplin et al., 2018) using a deep learning model retrained on exome data sequenced with the same protocol as was used to sequence the UKB 200K samples. Variant calls were restricted to the exome capture region and the 100 base-pairs flanking each capture target, resulting in a gVCF (genomic VCF) for each sample containing all variant genotypes and compressed representations of reference regions without called variant genotypes. All gVCFs were then merged with GLnexus 1.2.6 and the default ‘DeepVariantWES’ parameters (M. Lin et al., 2018; Yun et al., 2020).

### Variant summary & Annotation

A set of QC metrics were applied to the variants included in Table1. Variants that passed the following criteria are included: individual and variant missingness <10%, Hardy Weinberg Equilibrium p-value>10^-15, minimum read coverage depth of 7 for SNPs and 10 for indels, at least one sample per site passed the allele balance threshold > 0.15 for SNPs and 0.20 for indels. Variants were annotated using SnpEff (Cingolani et al., 2012) based on Ensembl 85 gene model. Transcript aware annotations are derived using protein coding transcripts with defined start and stop coordinates.

### Canonical transcript annotation

Transcripts with a “MANE Select v0.9” tag were obtained from Ensembl and were used to identify the canonical transcript of a gene. MANE annotation represents one high-quality transcript per protein-coding gene that is well-supported by experimental data and represents the biology of the gene. In the absence of a “MANE” annotation for a gene, APPRIS (Rodriguez et al., 2018) and Ensembl canonical annotation tags were used to define the canonical transcript set. Ensembl canonical annotation is obtained using the definition provided at http://uswest.ensembl.org/Help/Glossary

### LOF annotation

While sequence-based annotation of LOF variants is straightforward, identifying true LOF events is not. We subset the predicted LOF variants to “strict LOF” category using the following criteria: 1) Excluded UTR splice variants, start lost and stop lost variants; 2) Excluded variants that are present in the last 5% of the protein and are predicted to escape nonsense-mediated decay; 3) Included only variants that affect the canonical transcript.

## UKB-ESC Research Team

^**1**^**Bristol Myers Squibb**

Oleg Moiseyenko,Carlos Rios, Saurabh Saha

^**2**^**Regeneron Pharmaceuticals Inc**.

Goncalo Abecasis, Nilanjana Banerjee, Christina Beechert, Boris Boutkov, Michael Cantor, Giovanni Coppola, Aris Economides, Gisu Eom, Caitlin Forsythe, Erin D. Fuller, Zhenhua Gu, Lukas Habegger, Marcus B. Jones, Rouel Lanche, Michael Lattari, Michelle LeBlanc, Dadong Li, Luca A. Lotta, Kia Manoochehri, Adam J. Mansfield, Evan K. Maxwell, Jason Mighty, Mrunali Nafde, Sean O’Keeffe, Max Orelus, Maria Sotiropoulos Padilla, Razvan Panea, Tommy Polanco, Manasi Pradhan, Ayesha Rasool, Thomas D. Schleicher, Deepika Sharma, Alan Shuldiner, Jeffrey C. Staples, Cristopher V. Van Hout, Louis Widom, Sarah E. Wolf

^**3**^**Biogen Inc**.

Sally John, Chia-Yen Chen, David Sexton, Varant Kupelian, Eric Marshall, Timothy Swan, Susan Eaton, Jimmy Z. Liu, Stephanie Loomis, Megan Jensen, Saranya Duraisamy

^**4**^**Takeda Pharmaceutical Company Ltd**

Jason Tetrault, David Merberg, Sunita Badola

^**5**^**Abbvie Inc**.

Mark Reppell, Jason Grundstad, Xiuwen Zheng

^**6**^**Alnylam Pharmaceuticals**

Aimee M. Deaton, Margaret M. Parker, Lucas D. Ward, Alexander O. Flynn-Carroll

^**7**^**AstraZeneca**

Caroline Austin (Business Development); Ruth March (Precision Medicine & Biosamples); Menelas N. Pangalos (BioPharmaceuticals R&D); Adam Platt (Translational Science & Experimental Medicine, Research and Early Development, Respiratory and Immunology); Mike Snowden (Discovery Sciences); Athena Matakidou, Sebastian Wasilewski, Quanli Wang, Sri Deevi, Keren Carss, Katherine Smith (Centre for Genomics Research, Discovery Sciences, BioPharmaceuticals R&D)

^**8**^**Pfizer, Inc**.

Morten Sogaard, Xinli Hu, Xing Chen, Zhan Ye

**Table S1:** Approved drugs with targets supported by human genetics evidence.

| Gene Symbol | Gene Description | Generic drug name | drug type | mode of action | indication for drug | References |
| --- | --- | --- | --- | --- | --- | --- |
| ABCC8 | ATP binding cassette subfamily C member 8 | sulphonylureas | small molecule | inhibitor | diabetes | (Mahajan et al., 2018; Nestorowicz et al., 1996; Otonkoski et al., 1999; Thomas et al., 1996) |
| KCNJ11 | potassium voltage-gated channel subfamily J member 11 | sulphonylureas | small molecule | inhibitor | diabetes | (Flannick et al., 2019; Mahajan et al., 2018; A. P. Morris et al., 2012; Xue et al., 2018) |
| ADRB1 | beta1-adrenergic receptor | metoprolol | small molecule | inhibitor | hypertension | (Giri et al., 2019; Hoffmann et al., 2017; Sung et al., 2018; Takeuchi et al., 2018) |
| APOB | apolipoprotein B | mipomersen | antisense therapeutic | inhibitor | familial hypercholesterolemia | (Pullinger et al., 1995; Rabès et al., 1997; Soria et al., 1989; Van Der Graaf et al., 2011) |
| AVPR2 | arginine vasopressin receptor 2 | vasopressin/desmopressin | hormone | agonist | diabetes insipidus | (Bichet et al., 1994; Carroll et al., 2006; Pasel et al., 2000; Rosenthal et al., 1992) |
| CASR | calcium-sensing receptor | cinacalcet | small molecule | agonist | hypercalcemia | (Bai et al., 1996; Chou et al., 1995; Cole et al., 2009; Nissen et al., 2007; Zajickova et al., 2007) |
| CCR5 | C-C chemokine receptor type 5 | maraviroc | small molecule | inhibitor | HIV infection | (Carrington et al., 1997; Novembre et al., 2005; Quillent et al., 1998; Sullivan et al., 2001; Zimmerman et al., 1997) |
| CTLA4 | cytotoxic T-lymphocyte associated protein 4 | abatacept | protein replacement therapy | agonist | rheumatoid arthritis | (Carrington et al., 1997; Novembre et al., 2005; Quillent et al., 1998; Sullivan et al., 2001; Zimmerman et al., 1997) |
| DRD2 | dopamine receptor D2 | risperidone | small molecule | inhibitor | schizophrenia | (Li et al., 2017; Pardiñas et al., 2018; Ripke et al., 2014) |
| EPOR | erythropoietin receptor | Recombinant erythropoietin | hormone | agonist | anemia | (Kichaev et al., 2019; Xiong et al., 2015) |
| ESR1 | estrogen receptor alpha | estrogens | hormone | agonist | osteoporosis | (J. A. Morris et al., 2019) |
| F10 | factor X | rivaroxaban | small molecule | inhibitor | thrombosis | (Messier et al., 1996; Watzke et al., 1990) |
| F2 | thrombin | dabigatran | peptide mimetic | inhibitor | thrombosis | (Hinds et al., 2016) |
| FSHR | follicle-stimulating hormone receptor | recombinant FSH | hormone | agonist | infertility associated with PCOS | (Aittomäki et al., 1995) |
| GHR | growth hormone receptor | recombinant GH | hormone | agonist | growth stature | (Ayling et al., 1997; Goddard et al., 1995) |
| HMGCR | HMG-CoA reductase | atorvastatin | small molecule | inhibitor | hypercholesterolemia | (Burkhardt et al., 2008; Kathiresan et al., |
|  |  |  |  |  |  | 2008) |
| IL12B | interleukin-12 | ustekinumab | monoclonal antibody | inhibitor | psoriasis | (Cargill et al., 2007) |
| IL6R | interleukin-6 receptor | tocilizumab | monoclonal antibody | inhibitor | rheumatoid arthritis | (Eyre et al., 2012; Lauffer et al., 2019; Okada et al., 2014) |
| INS | insulin | recombinant insulin | hormone | agonist | diabetes | (Colombo et al., 2008; Edgill et al., 2008; Støy et al., 2007) |
| KCNH2 | hERG | amiodarone | small molecule | inhibitor | atrial fibrillation | (Sinner et al., 2008) |
| MTTP | microsomal triglyceride transfer protein, MTP | lomitapide | small molecule | inhibitor | familial hypercholesterolemia | (Khatun et al., 2013) |
| NPC1L1 | NPC1 like intracellular cholesterol transporter 1 | ezetimibe | small molecule | inhibitor | Hypercholesterolemia | (Stitzel et al., 2014) |
| P2RY12 | P2Y receptor 12 | clopidogrel | small molecule | inhibitor | "atherosclerosis, stroke" | (Cattaneo, 2011) |
| PCSK9 | PCSK9 | alirocumab, evolocumab | monoclonal antibody | inhibitor | hypercholesterolemia | (Abifadel et al., 2003; Cohen et al., 2005) |
| PNLIP | pancreatic lipase | orlistat | small molecule | inhibitor | obesity | (Winkler et al., 2015) |
| PPARG | peroxisome proliferator activated receptor gamma | rosiglitazone | small molecule | inhibitor | diabetes | (Zhao et al., 2017) |
| TNFSF11 | TNF superfamily member 11 | denosumab | monoclonal antibody | inhibitor | osteoporosis | (Estrada et al., 2012) |
| SOST | sclerostin | romosozumab | monoclonal antibody | inhibitor | osteoporosis | (Estrada et al., 2012; Hamersma et al., 2003; Stykars dottir et al., 2009, 2016) |
| VEGFA | vascular endothelial growth factor A | afibercept | recombinant protein | decoy receptor | AMD; Macular Edema Following Retinal Vein Occlusion; Diabetic Macular Edema; Diabetic Retinopathy | (J. M. Lin et al., 2008; Yu et al., 2011) |
| VEGFA | vascular endothelial growth factor A | ranimizumab; bevacizumab; brolucizumab | monoclonal antibody | inhibitor | AMD | (J. M. Lin et al., 2008; Yu et al., 2011) |
| VEGFA | vascular endothelial growth factor A | pegaptanib | small molecule | inhibitor | AMD | (J. M. Lin et al., 2008; Yu et al., 2011) |
| PPARA | peroxisome proliferator activated receptor alp | clofibrate; gemfibrozil; ciprofibrate; bezafibrate; and fenofibrate | small molecule | agonist | hypercholesterolemia | (Willer et al., 2013) |
| BCL2 | BCL2, apoptosis regulator | venetoclax | small molecule | inhibitor | chronic lymphocytic leukemia | (Berndt et al., 2013) |
| CD80 | CD80 molecule | belimumab | monoclonal antibody | inhibitor | systemic lupus erythematosus | (W. Yang et al., 2013) |
| SMN2 | survival of motor neuron 2, centromeric | nusinersen | antisense therapeutic | inhibitor | spinal muscular atrophy | (Aartsma-Rus, 2018) |
| LDLR | low density lipoprotein receptor | statins | small molecule |  | hyperlipidaemia | (Rader et al., 2003) |
| SRD5A2 | steroid 5 alpha-reductase 2 | finasteride | small molecule | inhibitor | benign prostrate hyperplasia | (Andersson et al., 1991; Imperato-McGinley et al., 1974; Wood & Rittmaster, 1994) |

